# PUBLIC HOSPITALS PREPAREDNESS IN THE PROVISION OF BREAST AND CERVICAL CANCER SERVICES IN BUSIA AND TRANS-NZOIA COUNTIES IN KENYA

**DOI:** 10.1101/2021.05.24.21257702

**Authors:** Peter Itsura, Grace W. Mwaura, Obed K. Limo, Gerald O. Lwande, Kenneth Too, Richard Mugo, Ann W. Mwangi, Nicholas Kirui, Philip Tonui, Gladwell Gathecha, Mary Nyangasi, Jemimah Kamano, Wilson Aruasa

**Affiliations:** Academic Model Providing Access to Healthcare (AMPATH), Eldoret-Kenya; Moi University School of Medicine, Eldoret-Kenya; Moi Teaching and Referral Hospital, Eldoret-Kenya; Ministry of Health, Division of Non-Communicable Diseases - Kenya

**Keywords:** Cervical Cancer, Breast Cancer, Preparedness, Service Provision Assessment (SPA), Public Healthcare Facilities, Western Kenya, Busia, Trans Nzoia

## Abstract

Cancer is a major cause of morbidity and mortality globally. This study aimed to determine the preparedness of public health care facilities in the provision of breast and cervical cancer services by assessing healthcare provider’s knowledge on risk factors, screening, symptoms, diagnosis and treatment of the diseases as well as availability of medical equipment required in their management in Western Kenya. This was achieved by using a cross-sectional service provision assessment (SPA) baseline survey technique in Busia and Trans-Nzoia counties of Western Kenya between October and December 2018. Data was collected using an interviewer-assisted questionnaires from healthcare workers, while a structured facility questionnaire was used to assess the level of preparedness of the selected public healthcare facilities. We enrolled 73 healthcare workers 37 (50.6%) of whom were nurses, followed by clinical officers and medical officers. The highest proportion of knowledge on risk factors and screening of breast and cervical cancer was reported among medical officers and consultant physicians while nurses scored highly on the symptoms of breast and cervical cancer. The medical equipment required for breast and cervical cancer screening and diagnosis were found in most healthcare facilities; however, none of them had core-biopsy needles. A single LEEP equipment was found in a health center within Trans Nzoia County while two LEEP equipment were stationed at the Busia county hospital. We report a below average level of knowledge on breast and cervical cancer among the healthcare workers at public healthcare facilities in both Busia and Trans Nzoia counties. There was a disparity in the distribution and quantity of priority medical equipment for the screening, diagnosis and treatment of breast and cervical cancer in the two county hospitals. There is need for capacity building among healthcare workers as well as standardization in the distribution and utilization of these medical equipment.

## Introduction

Cancer is a major cause of morbidity and mortality across the globe. According to the International Agency for Research on Cancer global cancer observatory report [1], approximately 1 in 5 people develop cancer during their lifetime. Of these, 1 in 8 men and 1 in 11 women die from the disease. With a rising proportion of the ageing populations globally, coupled with lifestyle changes, the prevalence of cancer is on the rise. The most common forms of cancer among women globally are breast, colorectal, lung, cervical, and thyroid cancers [2]. The likelihood of breast cancer is 1 in 4 cancers diagnosed among women globally [1,2]. In Africa the incidence for breast cancer in 2020 was estimated at 16.8% followed by cervical cancer at 10.6% [3]. In Kenya, the incidence of breast cancer in 2020 was estimated at 16.1% followed by second placed cervical cancer at 12.4% of all women diagnosed with cancer [2]. With a greater adoption of a more Westernized lifestyle (especially in diet, reproductive health and reduced physical activity) in Kenya and many other sub-Saharan African countries, the incidence of breast and cervical cancer has seen a steady rise [4]. These low- and medium-income countries (LMIC) have been shown to bear a greater burden of various forms of cancer despite advancements in screening, diagnosis and management options [5]. To adequately address the disease burden attributed to these two cancers, there is need to have effective and adequately prepared health service delivery systems. The level of preparedness and effectiveness of these health systems can be assessed using multiple dimensions to assess the progress in addressing the disease burden. Although various tools could be used in this assessment, a Service Provision Assessment (SPA) survey could be used to provide a comprehensive understanding of the health service delivery system in a particular country [6]. This is because they support the strengthening of health systems in developing countries by collecting information on the availability of different resources based in the health facilities within a country. The SPA survey describes the facility-types with the ability to offer specific health services. It defines the infrastructural and human resource preparedness to offer the said services in the health facilities [6]. Furthermore, it assesses adherence to the set quality standards for healthcare service provision and whether the clients and service providers are content with the service delivery environment.

The World Health Organization’s Global action plan for the prevention and control of noncommunicable diseases 2013-2020 targeted a 25% reduction in risk of premature death attributed to non-communicable diseases including cancer by 2025 [7]. Furthermore, the United Nations Agenda for Sustainable Development recommends a one-third reduction in the proportion of premature death associated with non-communicable diseases such as cancer by the year 2030 using multiple approaches such as prevention, treatment and promotion of mental health and well-being [8]. The public and private health care service delivery institutions should have an 80% availability of the affordable basic technologies and essential medicines for the management of these noncommunicable diseases. The report [7] recommends periodic needs assessment and evaluation of resource need such as workforce, institutional and research capacity to address the non-communicable diseases. The eight key indicators include a comprehensive range of health services provided to the target population, the services should be directly and permanently accessible without barriers of entry, they should adequately cover the defined target population and provide continuity of care across the network of services, health conditions, levels of care, and over the life cycle. The quality of these services should be high as evidenced by their effectiveness and safety as well as being organized around the person and not the disease or the financing mechanism. They should be properly coordinated at the local level and across the different types of providers, types of care and levels of service delivery with adequate accountability and efficiency [7].

In Kenya, the healthcare service delivery system is stratified into four tiers as [9]: Tier 1 (Community Health Services), Tier 2 (primary healthcare facilities such as dispensaries and health centers), Tier 3 (secondary referral facilities such as sub county and county hospitals) and Tier 4 (Tertiary facilities such as national referral hospitals). The primary care for cancer involves prevention of risk factors and screening, enhancing health promotion through public awareness, social mobilization and community engagement which are carried out in all the four tiers of healthcare service provision. Specifically, screening services begin from dispensaries and health centers towards higher tier facilities [10]. On the hand, cancer diagnosis and treatment is predominantly offered in secondary and tertiary referral facilities [10]. According Kenya’s Cancer Policy for 2019-2030, it is argued that nearly half (40%) of the cancers can be prevented by mitigating the risk factors and implementing already established evidence-based prevention strategies [9]. The common risk factors include being overweight or obesity (27.9%), inadequate consumption of fruits and vegetables (94%), tobacco and alcohol use at 13.3% and 19.3% respectively [9]. It is estimated that many women harbor human papilloma virus (HPV) types 18 (63.1%) followed by 16 (9.1%) strains that is attributed to invasive cervical cancer [11]. However, it has been previously reported that there is a low awareness of cancer and its risk factors in Kenya [12] both in the general population and among health care service providers [13,14]. Although the National Cancer Screening Guidelines have been disseminated and outline the key cancers for screening as well as the modalities for screening by level of care, there are gaps in the implementation of these guidelines for prevention and screening of cancer. The guidelines recommend screening for women 25-49 years with Human Papillomavirus (HPV) testing as the gold standard method for cervical cancer screening although Visual Inspection with Acetic Acid and Cytology are also recommended where HPV is not available. For breast cancer, mammography is the recommended screening method for women from 40-74 years of age. This necessitated a local study to determine healthcare workers knowledge on screening, diagnosis and treatment of breast and cervical cancer.

The Kenya Government set medium term plans for the implementation of vision 2030. The current third medium-term plan (2018-2022) set the universal healthcare coverage (UHC) policy to ensure the provision of high quality and affordable healthcare for all its citizens [15]. To achieve this, the government noted an insufficiency of medical equipment in both county and national hospitals. The government rolled-out the managed equipment scheme (MES) as a private-public partnership (PPP) between the government and private medical equipment service providers [16]. For the diagnosis and management of cancer, the Kenyan government supplied medical imaging equipment such as X-rays, Ultrasounds, CT-Scans, digital mammography and MRI machines through MES in all the 47 counties [9]. Both Busia and Kitale County referral hospitals received a digital mammography machine each for screening and early diagnosis of breast cancer. Although this project was noble, it was noted that the managed equipment scheme was not properly implemented due to inadequate personnel [17] to operate them and the backbone supporting infrastructure such as availability of electricity [16]. Because of this challenge, this study aimed to assess the preparedness of health care facilities in provision of Breast and Cervical Cancer services in Trans Nzoia and Busia Counties in Kenya. It established the level of healthcare workers knowledge on the screening, diagnosis and treatment of breast and cervical cancers as well the availability of medical equipment and supplies for screening, diagnosis and management of the two diseases in selected two counties of Western Kenya.

## Methods

This cross-sectional study was conducted in Busia and Trans Nzoia counties in Western Kenya, whose breast and cervical cancer oncology programmes are supported by the Academic Model Providing Access To Healthcare – AMPATH [18,19]. Trans Nzoia county is in the former Rift Valley Province of Kenya, it borders Bungoma county to the west, Uasin Gishu and Kakamega counties to the south, Elgeyo Marakwet county to the east and the Republic of Uganda to the Northwest. It covers a geographical area of 2495.5 km^2^ with a population of 956,559 of whom 49.7% are male. Approximately half of the population in Trans Nzoia county is aged 18 to 64 years with 2.8% aged more than 65 years and an average life expectancy of 60.5 years which is lower than the national average at 63.4 years. Busia county borders Kakamega county to the east, Bungoma county to the north, Siaya county to the south and Uganda to the west. It has a geographical area of 1628.4 Kilometer Squared (Km^2^) with a population of 953,337 of whom 52% are above 18 years of age and a low life expectancy of 47 years. The two counties have been reported to have a significantly high prevalence of breast and cervical cancer in Kenya.

The study adopted service provision assessment (SPA) survey technique to assess the healthcare providers level of knowledge on breast and cervical cancer as well as determine the availability of medical equipment as per the WHO priority list between September to December 2018. Healthcare workers heading oncology programs in the selected healthcare facilities were purposively sampled to saturation prior to obtaining written informed consents and being enrolled into the study. An interviewer assisted questionnaire was used to collect socio-demographic, knowledge of breast and cervical cancer risk factors, screening strategies, symptoms, diagnosis and treatment of the disease among the healthcare workers enrolled. Healthcare facilities were selected using a census approach and stratified by level of care offered. A structured facility questionnaire was used to explore the selected institutions’ preparedness to screen, diagnose and treat breast and cervical cancer. Specifically, the facility questionnaire assessed level of care provided and availability of specific equipment as per the WHO priority list. Data collection was done by trained research assistants who administered questionnaires which had been pre-programmed into the Research Electronic Data capture (REDCap) database mobile application which allowed for direct data entry, daily cleaning and validation by the team. Healthcare workers responses from the questionnaire were scored (with a point for every correct response) to assess knowledge on breast and cervical cancer and weighted means computed with their corresponding standard deviations. Descriptive statistical analysis of frequency with corresponding proportions were used to compare the availability of medical equipment in the two counties under review. All analysis were computed using standard statistical (STATA) version 15. This study received ethical approval from the Institutional Research and Ethics Committee (IREC) of MTRH/Moi University (approval number 0002090). This was followed by a research permit from the National Commission for Science technology and Innovation (NACOSTI) of Kenya (approval number NACOSTI/P/18/74238/24329) and the two County Health Management Teams (CHMTs). Data confidentiality was ensured by deidentifying participants responses while privacy was enhanced through used of password protected database and limited access to the datasets.

## Results

### Healthcare workers’ knowledge on breast and cervical cancer

This study enrolled 73 healthcare service providers from two counties (Busia and Trans Nzoia) in Western Kenya. They were stratified based on the level of training as either clinical officers (m=29), nurses (n=37), medical officers or consultants (n=7) and assessed on their knowledge of risk factors, symptoms, screening, diagnosis and treatment of breast and cervical cancer. The mean number of correct answers were scored based on the probable total correct responses.

On Breast cancer, there were varying responses on the probable risk factors for breast cancer. Nearly half (41%) of the clinical officers thought that a history of alcohol consumption was a major risk factor followed by using contraceptive pills (31%). However, 35% of the nurses described having a first child at an older age followed by alcohol consumption (24%) as the major risk factors for breast cancer. Similarly, more than half of medical officers and consulting clinicians considered alcohol consumption as a risk factor for breast cancer. All the healthcare workers enrolled described mastectomy alone as the treatment for breast cancer. There were higher levels of knowledge on breast cancer risk factors and screening among medical officers or consultant physicians compared to clinical officers and nurses. However, nurses appeared to have a higher mean knowledge score on the symptoms for breast cancer.

Regarding cervical cancer, the overall mean knowledge score was highest among medical officers or consultant and lowest among nurses across all domains assessed namely: risk factors, screening, diagnosis and treatment). Notably, nurses had more knowledge on symptoms of both cervical and breast cancers compared to other cadres (Table 1).

**Table 1:**
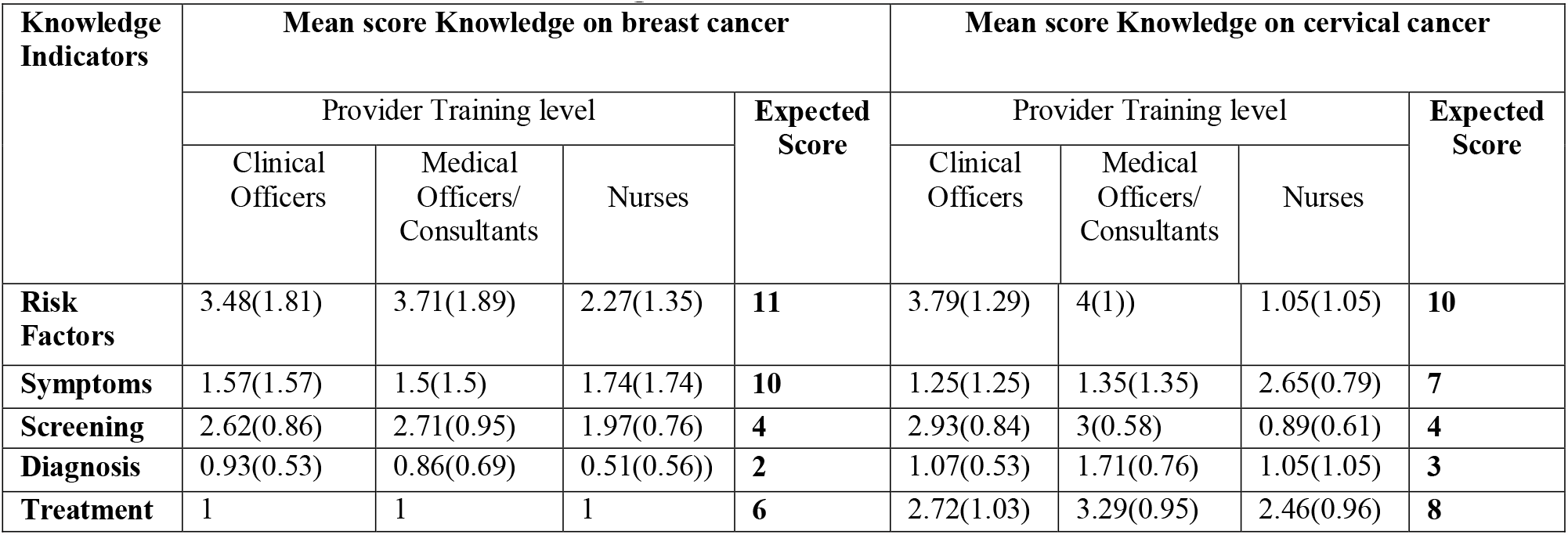
Level of Provider Knowledge.

| Knowledge Indicators | Mean score Knowledge on breast cancer |  |  |  | Mean score Knowledge on cervical cancer |  |  |  |
| --- | --- | --- | --- | --- | --- | --- | --- | --- |
|  | Provider Training level |  |  | Expected Score | Provider Training level |  |  | Expected Score |
|  | Clinical Officers | Medical Officers/ Consultants | Nurses |  | Clinical Officers | Medical Officers/ Consultants | Nurses |  |
| **Risk Factors** | 3.48(1.81) | 3.71(1.89) | 2.27(1.35) | **11** | 3.79(1.29) | 4(1)) | 1.05(1.05) | **10** |
| **Symptoms** | 1.57(1.57) | 1.5(1.5) | 1.74(1.74) | **10** | 1.25(1.25) | 1.35(1.35) | 2.65(0.79) | **7** |
| **Screening** | 2.62(0.86) | 2.71(0.95) | 1.97(0.76) | **4** | 2.93(0.84) | 3(0.58) | 0.89(0.61) | **4** |
| **Diagnosis** | 0.93(0.53) | 0.86(0.69) | 0.51(0.56)) | **2** | 1.07(0.53) | 1.71(0.76) | 1.05(1.05) | **3** |
| **Treatment** | 1 | 1 | 1 | **6** | 2.72(1.03) | 3.29(0.95) | 2.46(0.96) | **8** |

### Medical equipment distribution

During the initial baseline survey conducted in both counties (Busia and Trans Nzoia), it was noted that Busia county only had 6 ultrasound machines available for breast cancer screening and with half of them (3) in sub-county hospitals, one in a health center while the county referral hospital had two available. It was striking that 35 (87.5%) of the 40 healthcare facilities in the county did not have a single ultrasound machine. Furthermore, x-ray equipment that would support in further screening was only available in 5 (12.5%) healthcare facilities within Busia County, with the county hospital and one-subcounty having one and two x-ray equipment respectively. For cervical cancer screening, speculum availability was nearly evenly distributed in the healthcare facilities in Busia County with 25% (n=10) of the facilities having between 6-10 speculums while 22.5% (n=9) having more than 21. Gynecological beds were only available in 12 (30%) of all the healthcare facilities assessed in Busia county with most of the gynecological beds being concentrated in County and sub-County hospitals. No public healthcare facility in Busia county had a biopsy needle for breast and cancer diagnosis while sterilization equipment were found in 90% of all the public healthcare facilities. There were seven (17.5%) had cryotherapy equipment available in the public healthcare facilities in Busia County, however nearly all (97.5%) of the healthcare facilities did not have a LEEP equipment for the management of cervical lesions with the only available ones (n=2) being in Busia County referral hospital (Table 2).

**Table 2:** Medical equipment availability by healthcare facility’s level of care in Busia County.

| Variable name | Value | Total<br>(N=40) | Sub County<br>n=7 (18%) | Health Centre<br>n=10 (25%) | Dispensary<br>n=22 (55%) | County Referral<br>n=1 (2%) |
| --- | --- | --- | --- | --- | --- | --- |
| **Ultrasounds machines** |  |  |  |  |  |  |
|  | 0 | 35 (87.5%) | 4 (57.1%) | 9 (90%) | 22 (100%) | 0 (0%) |
|  | 1 | 4 (10%) | 3 (42.9%) | 1 (10%) | 0 (0%) | 0 (0%) |
|  | 2 | 1 (2.5%) | 0 (0%) | 0 (0%) | 0 (0%) | 1 (100%) |
| **X-ray machine** |  |  |  |  |  |  |
|  | 0 | 35 (87.5%) | 3 (42.9%) | 10 (100%) | 22 (100%) | 0 (0%) |
|  | 1 | 3 (7.5%) | 3 (42.9%) | 0 (0%) | 0 (0%) | 0 (0%) |
|  | 2 | 1 (5%) | 0 (0%) | 0 (0%) | 0 (0%) | 1 (100%) |
| **Speculum available** |  |  |  |  |  |  |
|  | 0 | 1 (2.5%) | 0 (0%) | 0 (0%) | 1 (4.5%) | 0 (0%) |
|  | 1-5 | 10 (25%) | 0 (0%) | 2 (20%) | 8 (36.4%) | 0 (0%) |
|  | 6-10 | 10 (25%) | 1 (14.3%) | 2 (20%) | 7 (31.8%) | 0 (0%) |
|  | 11-15 | 7 (17.5%) | 4 (57.1%) | 2 (20%) | 1 (4.5%) | 0 (0%) |
|  | 16-20 | 3 (7.5%) | 1 (14.3%) | 0 (0%) | 1 (4.5%) | 1 (100%) |
|  | Above 21 | 9 (22.5%) | 1 (14.3%) | 4 (40%) | 4 (18.2%) | 0 (0%) |
| **Gynecological beds** |  |  |  |  |  |  |
|  | 0 | 28 (70%) | 4 (57.1%) | 6 (60%) | 18 (81.8%) | 0 (0%) |
|  | 1 | 6 (15%) | 0 (0%) | 3 (30%) | 3 (13.6%) | 0 (0%) |
|  | 2 | 6 (15%) | 3 (42.9%) | 1 (10%) | 1 (4.5%) | 1 (100%) |
| **Biopsy needles** |  |  |  |  |  |  |
|  | 0 | 40 (100%) | 7 (100%) | 10 (100%) | 22 (100%) | 1 (100%) |
| **Sterilization Equipment** |  |  |  |  |  |  |
|  | 0 | 4 (10%) | 0 (0%) | 0 (0%) | 4 (18.2%) | 0 (0%) |
|  | 1 | 23 (57.5%) | 3 (42.9%) | 6 (60%) | 14 (63.6%) | 0 (0%) |
|  | 2 | 10 (25%) | 3 (42.9%) | 4 (40%) | 2 (9.1%) | 1 (100%) |
|  | 3 | 2 (5%) | 0 (0%) | 0 (0%) | 2 (9.1%) | 0 (0%) |
|  | 4 | 1 (2.5%) | 1 (14.3%) | 0 (0%) | 0 (0%) | 0 (0%) |
| **Cryotherapy equipment** |  |  |  |  |  |  |
|  | 0 | 33 (82.5%) | 5 (71.4%) | 9 (90%) | 19 (86.4%) | 0 (0%) |
|  | 1 | 6 (15%) | 2 (28.6%) | 1 (10%) | 3 (13.6%) | 0 (0%) |
|  | 4 | 1 (2.5%) | 0 (0%) | 0 (0%) | 0 (0%) | 1 (100%) |
| **LEEP equipment** |  |  |  |  |  |  |
|  | 0 | 39 (97.5%) | 7 (100%) | 10 (100%) | 22 (100%) | 0 (0%) |
|  | 2 | 1 (2.5%) | 0 (0%) | 0 (0%) | 0 (0%) | 1 (100%) |

In Trans Nzoia County, there were 7 ultrasound machines serving 33 public healthcare facilities within the county for the screening of breast cancer. There was only one out of the six sub-county hospitals with an ultrasound machine, while 2 of the 15 health centers had 2 ultrasound machines each and the county referral hospital equally had 2 machines. This study noted that there were 9 x-ray machines that could support in cancer screening within Trans Nzoia County where a county and sub-county hospital having 2 each while a health center having three x-ray machines. One of the X-ray machines reported in the county hospital of Trans Nzoia was a digital mammogram. Most (36.4%; n=12) of the facilities assessed had a 1-5 speculum for cervical cancer screening followed by those with 6-10 speculum in 11 (33.3%) facilities. However, a single dispensary did not have any speculum. Public healthcare facilities in Trans Nzoia County had 44 gynecological beds distributed among 27 of the 33 facilities with most of the gynecological beds being concentrated in County (4) and sub-County (12) hospitals. Similar to Busia County, no public healthcare facility in Trans Nzoia County had a biopsy needle for cervical cancer diagnosis. However, only 4 (12.1%) of the facilities surveyed in Trans Nzoia County lacked a sterilization equipment. There were 13 (39.4%) cryotherapy equipment available in the public healthcare facilities within Trans Nzoia County, where majority (8) of these were in health centers. This study reports the availability of a single LEEP equipment stationed at a health center in Trans Nzoia County (Table 3).

**Table 3:**
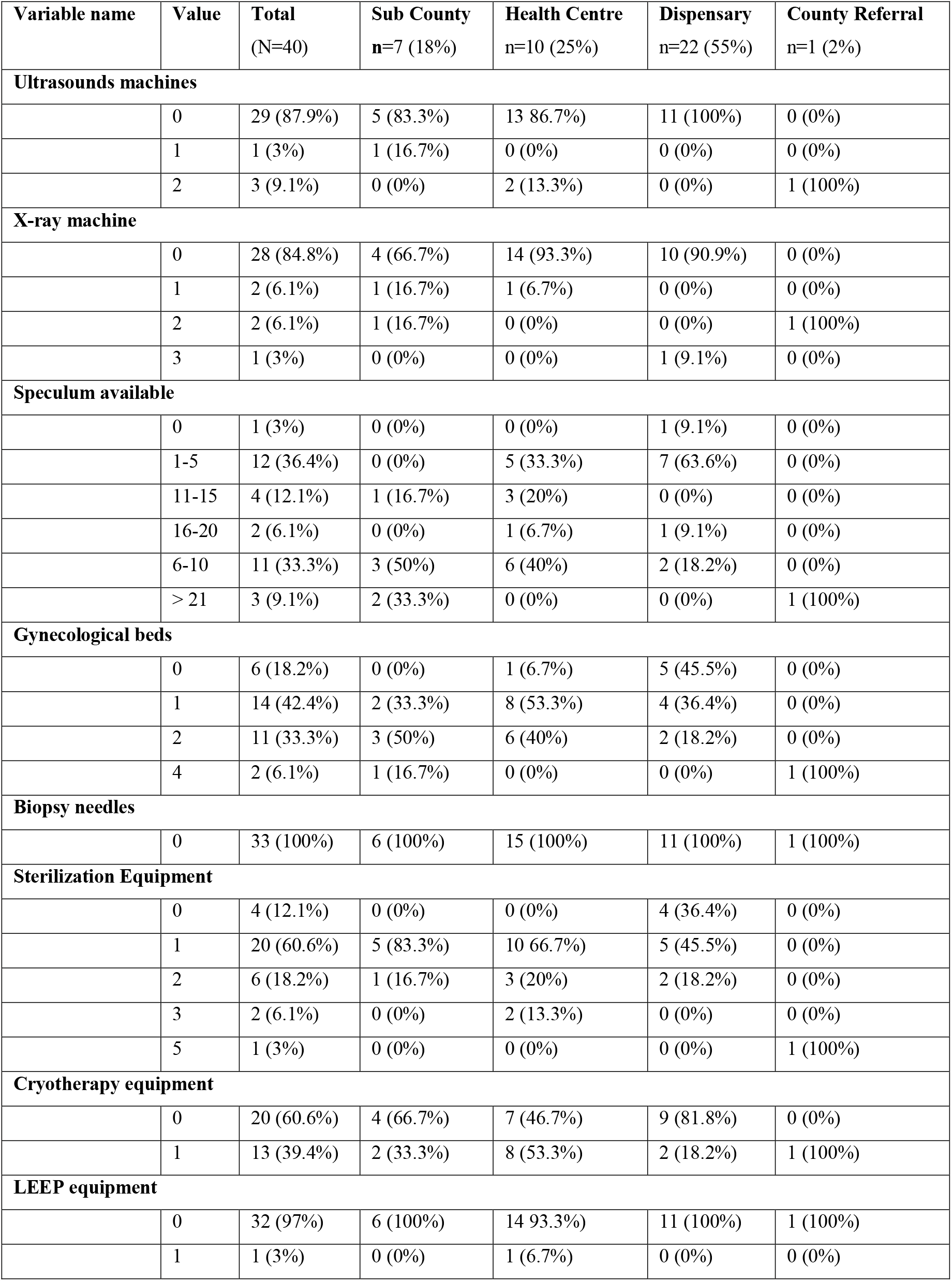
Medical equipment availability by healthcare facility’s level of care in Trans Nzoia County.

| Variable name | Value | Total<br>(N=40) | Sub County<br>n=7 (18%) | Health Centre<br>n=10 (25%) | Dispensary<br>n=22 (55%) | County Referral<br>n=1 (2%) |
| --- | --- | --- | --- | --- | --- | --- |
| **Ultrasounds machines** |  |  |  |  |  |  |
|  | 0 | 29 (87.9%) | 5 (83.3%) | 13 86.7%) | 11 (100%) | 0 (0%) |
|  | 1 | 1 (3%) | 1 (16.7%) | 0 (0%) | 0 (0%) | 0 (0%) |
|  | 2 | 3 (9.1%) | 0 (0%) | 2 (13.3%) | 0 (0%) | 1 (100%) |
| **X-ray machine** |  |  |  |  |  |  |
|  | 0 | 28 (84.8%) | 4 (66.7%) | 14 (93.3%) | 10 (90.9%) | 0 (0%) |
|  | 1 | 2 (6.1%) | 1 (16.7%) | 1 (6.7%) | 0 (0%) | 0 (0%) |
|  | 2 | 2 (6.1%) | 1 (16.7%) | 0 (0%) | 0 (0%) | 1 (100%) |
|  | 3 | 1 (3%) | 0 (0%) | 0 (0%) | 1 (9.1%) | 0 (0%) |
| **Speculum available** |  |  |  |  |  |  |
|  | 0 | 1 (3%) | 0 (0%) | 0 (0%) | 1 (9.1%) | 0 (0%) |
|  | 1-5 | 12 (36.4%) | 0 (0%) | 5 (33.3%) | 7 (63.6%) | 0 (0%) |
|  | 11-15 | 4 (12.1%) | 1 (16.7%) | 3 (20%) | 0 (0%) | 0 (0%) |
|  | 16-20 | 2 (6.1%) | 0 (0%) | 1 (6.7%) | 1 (9.1%) | 0 (0%) |
|  | 6-10 | 11 (33.3%) | 3 (50%) | 6 (40%) | 2 (18.2%) | 0 (0%) |
|  | > 21 | 3 (9.1%) | 2 (33.3%) | 0 (0%) | 0 (0%) | 1 (100%) |
| **Gynecological beds** |  |  |  |  |  |  |
|  | 0 | 6 (18.2%) | 0 (0%) | 1 (6.7%) | 5 (45.5%) | 0 (0%) |
|  | 1 | 14 (42.4%) | 2 (33.3%) | 8 (53.3%) | 4 (36.4%) | 0 (0%) |
|  | 2 | 11 (33.3%) | 3 (50%) | 6 (40%) | 2 (18.2%) | 0 (0%) |
|  | 4 | 2 (6.1%) | 1 (16.7%) | 0 (0%) | 0 (0%) | 1 (100%) |
| **Biopsy needles** |  |  |  |  |  |  |
|  | 0 | 33 (100%) | 6 (100%) | 15 (100%) | 11 (100%) | 1 (100%) |
| **Sterilization Equipment** |  |  |  |  |  |  |
|  | 0 | 4 (12.1%) | 0 (0%) | 0 (0%) | 4 (36.4%) | 0 (0%) |
|  | 1 | 20 (60.6%) | 5 (83.3%) | 10 66.7%) | 5 (45.5%) | 0 (0%) |
|  | 2 | 6 (18.2%) | 1 (16.7%) | 3 (20%) | 2 (18.2%) | 0 (0%) |
|  | 3 | 2 (6.1%) | 0 (0%) | 2 (13.3%) | 0 (0%) | 0 (0%) |
|  | 5 | 1 (3%) | 0 (0%) | 0 (0%) | 0 (0%) | 1 (100%) |
| **Cryotherapy equipment** |  |  |  |  |  |  |
|  | 0 | 20 (60.6%) | 4 (66.7%) | 7 (46.7%) | 9 (81.8%) | 0 (0%) |
|  | 1 | 13 (39.4%) | 2 (33.3%) | 8 (53.3%) | 2 (18.2%) | 1 (100%) |
| **LEEP equipment** |  |  |  |  |  |  |
|  | 0 | 32 (97%) | 6 (100%) | 14 93.3%) | 11 (100%) | 1 (100%) |
|  | 1 | 1 (3%) | 0 (0%) | 1 (6.7%) | 0 (0%) | 0 (0%) |

## Discussion

### Knowledge on breast and cervical cancer

In this study there was a low level of knowledge among health care providers on the risk factors, symptoms, screening, diagnosis and treatment of both breast and cervical cancer. This finding is consistent with a previous study conducted in Kenya [20] which found low level of knowledge among healthcare workers on breast and cervical cancer. This finding was across various cadres and level of training of the healthcare workers. This low level of knowledge could directly influence the kind of care patients with both cancers could receive in public healthcare facilities located in the devolved governance units of Kenya. In a community study conducted in the coastal region of Kenya [12], the key barriers affecting access breast cancer care were lack of awareness on breast and cervical cancer risk factors, signs and symptoms among the community members enrolled. In addition to stigma towards patients seeking cervical cancer treatment and the overall negative attitudes towards patients with cancer, cultural beliefs and personal discomfort were some of the other reasons for given low access to cancer care. Among the health care workers who were interviewed, it was noted that they too had limited knowledge on breast cancer [12]. The Kenya Cancer Policy of 2019 to 2030 further acknowledges that there exist a knowledge-gap in the level of knowledge on various forms of cancer among human resources for health [9]. The Ministry of Health in Kenya has instituted various mechanisms to train more healthcare workers on by conducting capacity building activities and mentorship to primary care providers in counties by coordinating specialist outreach programs in collaboration with tertiary facilities. Furthermore, Kenya has initiated diploma, masters and oncology fellowship programs in addition to ongoing pre-service and in service training for primary care workers to strengthen cancer care service delivery [9,10].

Low levels on knowledge in cancer are not limited to healthcare workers practicing in public hospitals in Kenya. In Rwanda [21], it was reported that cancer care is mainly offered by physicians who work in collaboration with colleagues in USA and Canada through virtual tumor boards. The authors [21] further noted that in Rwanda, there are gaps in prevention and early detection of breast and cervical cancer despite introduction of mitigation measures such as breast surgery fellowship at the national military hospital.

### Medical Equipment

This study based its assessment on the availability and distribution of medical equipment for the management of cancer on the World Health Organization’s priority list of medical devices for cancer management of 2017 [22]. This priority list provides an implementation reference to be adopted based on the needs of the healthcare system in the specific country and serves as a standard by which we can assess preparedness to provide cancer care. The guideline stipulates the minimum requirements for prevention, diagnosis, treatment, follow up and palliative care for all cancers. For breast cancer prevention, physical examination through bimanual palpation of breasts and locoregional lymph nodes as well as mammography is recommended in low resource healthcare settings, while an MRI breast may be used in combination with mammography in certain high-risk patients. Among the 73 facilities assessed, none of them had an MRI machine to support breast cancer screening, however each county referral hospital had a digital mammogram. This finding is consistent to that reported in Rwanda [21] where MRI machines were available in one national public and one private hospital. The diagnosis of breast cancer involves triple assessment through a clinical breast examination by a trained health provider, breast biopsy and breast imaging using mammography, ultrasound or MRI as appropriate [23]. Biopsy sampling is usually performed for histopathological diagnosis and biomarker analysis of the sample. In the facilities assessed in this study, ultrasound services were available in 5 of the 40 public healthcare facilities in Busia County while in Trans Nzoia only 4 out of the 33 facilities in the county. However, none of these counties reported having biopsy needles to support histopathological diagnosis of breast cancer. This is because only a few county hospitals in Kenya have the capacity to offer histopathology services contributing to diagnostic delays [9]. Computerized tomography scanning of the chest as well as a chest x-Ray in low resource settings could be used in the staging of breast cancer. In this study, x-ray services were not available in many facilities in both Trans Nzoia and Busia counties with only 9 and 5 machines available respectively. The current cancer Policy in Kenya [9] as well as the Kenya Harmonized Health Facility Assessment 2018/2019 [24] highlight that the major challenge in adequate distribution of diagnostic equipment to healthcare institutions in Kenya is a mismatch between human resource capacity to operationalize the equipment and supplies. This could be a major reason for the low utilization of the already existing medical equipment. Furthermore, the policy notes a lack of clear policy guideline on distribution, placement, monitoring and evaluation of medical equipment. That is why this study noted a lack of consistency in the distribution of ultrasound and x-ray machines across the various tiers of the healthcare facilities in the counties reviewed. All these equipment availability challenges in both counties could be a major contributor to the late diagnosis of breast cancer currently being witnessed in Kenya [2,9,10].

The current cervical cancer screening and prevention strategies [11,25] involve preventive immunization against human papilloma virus (HPV) for girls, screening of pre-cancerous lesions using a speculum or vaginal examinations and inspection with acetic acid (VIA), conducting of a HPV test, cytology and treating of pre-cancerous lesions using cryotherapy and large loop excision of transformation (LEEP). In this study, most facilities assessed in Trans Nzoia had a minimum of 1-5 speculums to support screening and a single gynecological bed available, however only a single sub-county facility had LEEP and a cryotherapy equipment. In Busia county, most facilities had 11-15 specula available, with two LEEP equipment in the county hospital and five cryotherapy equipment. A biopsy needle is needle is needed in the definitive diagnosis of invasive breast cancer in the second care level [22]. However, no facility in both counties had a core biopsy needle. An abdominal ultrasound is often used in the staging of cervical cancer as well as chest x-ray. However, there were limited ultrasound and x-ray facilities in the studies sampled.

### Conclusions and Recommendations

We report a below average level of knowledge on breast and cervical cancer among the healthcare workers attending to patients in public healthcare facilities within both Busia and Trans Nzoia counties. Furthermore, there was a disparity in the distribution and quantity of priority medical equipment for the screening, diagnosis and treatment of breast and cervical cancer in the two county hospitals. From the findings of this study, there is need for targeted in-service training among nurses on breast and cervical cancer risk factors and screening modalities as they are the first contacts with suspected cancer patients and are also involved patient medical education. Advanced cancer diagnostic equipment such as core-biopsy needles should be provided in these public healthcare facilities to support breast cancer staging and risk assessment. There is need for specialized training of healthcare workers to reduce the mismatch between human resource capacity to operationalize the medical equipment and their supply. Finally, proper guidelines should be drafted in the distribution of medical equipment by the facility’s level of care and human resource availability.

## Data Availability

The data sets used during this study can be made available by the corresponding author on request.

## Acknowledgements

This research was supported with funds from Access Accelerated and by cooperative agreement between World Bank and Ministry of Health Kenya NCDs Division. The management of Moi Teaching and Referral Hospital (MTRH), Moi University and the Counties of Busia and Trans Nzoia for an implementation agreement that enabled the data collection in the selected public hospitals. We further acknowledge the study participants as well as AMPATH for providing technical support for this study.

## Authors’ contribution

All the authors equally contributed to the writing and final approval prior to submission.

